# ShapeMed-Knee: A Dataset and Neural Shape Model Benchmark for Modeling 3D Femurs

**DOI:** 10.1101/2024.05.06.24306965

**Authors:** Anthony A. Gatti, Louis Blankemeier, Dave Van Veen, Brian Hargreaves, Scott L. Delp, Garry E. Gold, Feliks Kogan, Akshay S. Chaudhari

## Abstract

Analyzing anatomic shapes of tissues and organs is pivotal for accurate disease diagnostics and clinical decision-making. One prominent disease that depends on anatomic shape analysis is osteoarthritis, which affects 30 million Americans. To advance osteoarthritis diagnostics and prognostics, we introduce *ShapeMed-Knee*, a 3D shape dataset with 9,376 high-resolution, medicalimaging-based 3D shapes of both femur bone and cartilage. Besides data, ShapeMed-Knee includes two benchmarks for assessing reconstruction accuracy and five clinical prediction tasks that assess the utility of learned shape representations. Leveraging ShapeMed-Knee, we develop and evaluate a novel hybrid explicit-implicit neural shape model which achieves up to 40% better reconstruction accuracy than a statistical shape model and two implicit neural shape models. Our hybrid models achieve state-of-the-art performance for preserving cartilage biomarkers (root mean squared error ≤ 0.05 vs. ≤ 0.07, 0.10, and 0.14). Our models are also the first to successfully predict localized structural features of osteoarthritis, outperforming shape models and convolutional neural networks applied to raw magnetic resonance images and segmentations (e.g., osteophyte size and localization 63% accuracy vs. 49-61%). The ShapeMed-Knee dataset provides medical evaluations to reconstruct multiple anatomic surfaces and embed meaningful disease-specific information. ShapeMed-Knee reduces barriers to applying 3D modeling in medicine, and our benchmarks highlight that advancements in 3D modeling can enhance the diagnosis and risk stratification for complex diseases. The dataset, code, and benchmarks are freely accessible.

## I. Introduction

Osteoarthritis (OA) is the leading cause of pain and disability in developed countries, impacting 30.8 million US adults [1] with an annual US cost of $180 billion [2]. OA affects all tissues in a joint, with emphasis on bone and cartilage. The majority of deep learning research in OA focuses on 2D convolutional neural networks (CNNs) applied to X-rays, 2D and 3D CNNs for segmentation of magnetic resonance images (MRI), and few studies using 3D CNNs for classification of MRIs [3], [4], [5], [6], [7], [8]. OA research largely focuses on X-rays due to the limitations of efficiently processing large 3D image volumes, however, X-rays are a 2D projection of the joint and are thus prone to parallax errors, particularly with repositioning [9].

Characterizing OA relies on medical imaging to discern the shape of anatomic tissues [10]. As OA progresses, osteophytes grow at the edges of cartilage, and cartilage is thinned. Radiographic OA diagnosis is primarily based on these shape features [10]. Beyond OA, shape analysis also serves as the basis for numerous health conditions and diagnoses. For example, shape modeling is crucial for diagnosis and treatment of craniosynostosis, a pediatric condition where skull bones fuse early, causing deformity and potential brain damage [11]. Numerous orthopedic conditions are related to bone shape; both gross shape [12], [13] and nuanced curvatures of joint articulations [14] are important for diagnosing, treating, and preventing disease.

Shape modeling provides an efficient way to analyze 3D anatomic data [15]. However, current shape models, and shape model research has limitations. Widely adopted statistical shape models (SSMs) require anatomic point matching, which is not guaranteed and, in disease, may not be possible. For example, osteophytes that form in OA are not present in healthy bones, and thus no true matching points exist. Once matching points are obtained, SSMs are typically fit using linear statistical representations, namely principal components analysis (PCA); shape features of disease are unlikely to be purely linear in nature. Applications of SSMs in medicine are typically used to identify gross features or predict disease in general [16], [17]; accurate quantification of specific, localized, biomarkers of disease are required for clinical applications. To advance shape analysis in medicine, we require benchmarks that assess clinically relevant reconstruction metrics, and whether a model can localize relevant disease features.

With our overarching objective to enable the advancement of medical domain-specific 3D modeling, we provide the following contributions (Fig. 1):

- We introduce ***ShapeMed-Knee***: a 3D anatomic dataset with 9,376 shapes, each including two interrelated objects (femur bone and cartilage). We publicly share segmentation masks, and 3D shapes^1^.
- We define seven medically relevant benchmark tasks with our ShapeMed-Knee dataset: surface reconstruction, cartilage biomarker calculation from reconstructions, disease diagnosis, localized disease staging, and future surgical event prediction.
- We develop hybrid explicit-implicit neural shape models (NSM) that outperform both SSMs and two implicit NSMs for bone and cartilage reconstruction (up to 20% lower average symmetric surface distance).
- We demonstrate that our hybrid NSM outperforms an SSM, two implicit NSMs, and CNNs in disease staging, disease diagnosis, and localization of specific features of disease.
- We show that interpolation in NSM latent space produces interpretable smooth interpolation of physical shape, clinical shape features, and clinical predictions.^2^
- We demonstrate precise control over localized disease features by interpolating latent space along classifier-fitted vectors, enabling targeted manipulations of disease characteristics.
- We publicly share our NSM model and the code used for training and inference^3^. A tutorial on how to download and use the data is provided^4^.

**Fig. 1.**
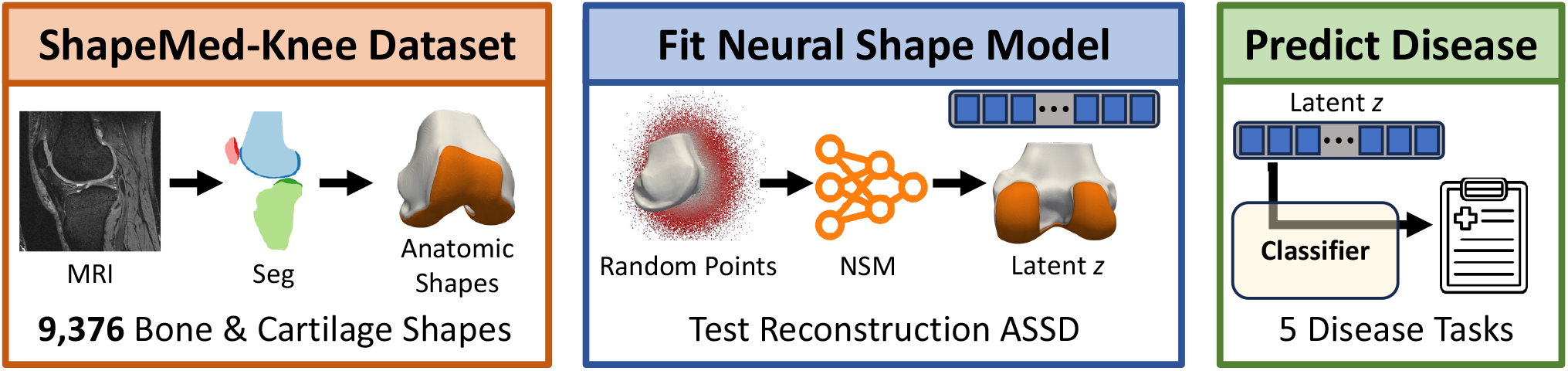
The ShapeMed-Knee dataset was created by segmenting and meshing 9,376 knee MRIs (orange box). We fit four shape models, three neural shape models (NSM) and one statistical shape model (SSM) to the ShapeMed-Knee training data and evaluated reconstruction tasks, including average symmetric surface distance (ASSD) (blue box). To test latent vectors ***z*** learned by the shape models, we train and evaluate classifiers for five clinical tasks (green box).

### II. Related Work

Neural representations have advanced computer graphics [18]. ShapeNet data has been central to the advancement of generative 3D shape models [19]. The recently proposed MedShapeNet is similar to ShapeNet, but includes 3D anatomic shapes with multiple inter-related tissues [20]. However, there still exists a gap in 3D anatomic models with curated disease-specific reconstruction metrics and clinical tasks; these data are needed to enable focused research that advances methods for quantifying anatomic shapes and understanding how these shapes influence health and disease.

### A. Generative Implicit Neural Representations

DeepSDF [21] and others [22], [23] first reported use of generative implicit neural representations. DeepSDF uses a multilayer perceptron (MLP) to generate shapes conditioned on a latent vector *z*. DeepSDF enables shape compression, interpolation, and completion from partial observations. Numerous DeepSDF advances have been proposed. Curriculum DeepSDF using curriculum learning [24]. Modulated Periodic Activations [25] combine two MLPs as a means of leveraging periodic (sinusoidal) activations [26].

To improve reconstruction of large scenes or fine details, instead of a single global *z*, a spatially localized *z* is input into the MLP [27], [28]. Hybrid explicit-implicit formulations generate localized *z* by leveraging the expressivity of CNNs [28], [29], [30], [31]. Both generative adversarial network and variational autoencoder (VAE) frameworks have been used in these hybrid explicit-implicit models [29], [30].

### B. Shape Modeling

Shape modeling has many important applications for biomedical data. In just the OA community, shape models have been used for automated segmentation [32], [33], disease prediction and staging [17], [34], [35], and generating synthetic data for physics-based simulations [14], [36]. Shape models have advanced understanding and treatment of conditions related to the heart, brain, skull, and bones, to name a few [37], [11], [38], [12], [13]. Improved shape modeling can benefit all of these areas, providing tangible benefits in understanding disease and improving patient health.

### C. Statistical Shape Models

Conventional SSMs use PCA to learn shape features. The main challenge with PCA-based SSMs for anatomical objects is the need for matching points at the same anatomical location on each object. Correspondence is typically obtained via nonrigid image registration of signed distance fields [32], or nonrigid point cloud registration [16], [14], [39]. To improve anatomic correspondence, registration features beyond XYZ coordinates, such as spectral coordinates or curvatures have been included [16], [40]. Registration is prone to failure in abnormal or diseased areas, which are typically the most important.

### D. Neural Shape Models

We refer to generative shape models in the medical domain as NSMs. There are only a handful of NSM applications. Amiranashvili et al. fit an occupancy NSM to anisotropic bone data showing occupancy-based methods can be trained and applied to undersampled anisotropic data. However, the occupancy NSMs still exhibit relatively large reconstruction errors (average symmetric surface distance (ASSD): 0.25-0.48mm) [41]. Jensen et al. fit a NSM by deforming points on a sphere using point-specific latent vectors. During training, a single latent vector was used for all points, while during inference, latents vary over the surface to increase expressivity. They showed better reconstruction than DeepSDF and improved segmentation results [42]. Ludke et al. used a neural flow deformer to fit a NSM by deforming coordinates from a template shape to the target, outperforming a conventional SSM in terms of surface reconstruction and simple OA classification [43].

Biomedical research demonstrates that implicit neural representations applied as NSMs improve anatomical reconstructions and image segmentation results and can encode basic clinical information. However, existing work represents only a single tissue at a time, uses relatively small samples of data (41-354 examples), and primarily focuses on surface reconstruction results rather than the quality of learned representations. Finally, biomedical approaches are challenging to compare as they use different datasets and downstream prediction tasks.

## III. Dataset & Evaluation

Data from this study is derived from the Osteoarthritis Initiative (OAI), a multi-center, longitudinal observational study of 4,796 men and women (45-79 years of age) with the goal of developing biomarkers of OA. The OAI collected patient clinical data, X-rays, and MRIs annually for 9 years. Important for the prediction tasks in this study, teams of expert radiologists were contracted to label acquired images for OA diagnosis, as well as standardized features of OA disease. We derive our dataset from the MR imaging data collected at the baseline time point and the radiologist evaluations from the baseline and all follow-up time points. The original OAI data, including the raw MRI data used in this study can be found at: https://nda.nih.gov/oai, all other data are available on Hugging Face^5^.

We used baseline data from the OAI, and thus each knee only appears once in the dataset. We used stratified random sampling to split the OAI baseline data into train/validation/test sets at the subject level; stratification was done at the subject level because right and left knees can be highly correlated and thus may be a form of data leakage. Splits were stratified over sex and clinical prediction tasks (IV) to ensure disease states and outcomes were equally represented. Due to the iterative and time-consuming nature of fitting the shape models during inference, a small validation set was used in this study (train: 67.5%, 3,233 people and 6,325 knees; validation: 2.5%, 74 people and 141 knees; test: 30.0%, 1,481 people and 2,910 knees). Tab. I contains an overview of the amount of data available for each task.

**TABLE I.** Amount of data for each evaluation task. *Subjects* is the number of individuals in each data split. *Recon* is the number of 3D models; Recon is at most **2 *×*** Subjects (one right, one left). The remaining rows indicate the number of knees that included the clinical outcome needed for the relevant prediction task, the max is the number of 3D models (Recon). KL = Kellgren Lawrence grade / osteoarthritis staging; OA = osteoarthritis (diagnosis) ; MOAKS = MRI Osteoarthritis Knee Score; (O) = osteophyte; (C) = cartilage hole or cartilage thinning; Future OA = future osteoarthritis; Future KR = future knee replacement

| Task | Train | Val | Test | Total |
| --- | --- | --- | --- | --- |
| Subjects | 3,233 | 74 | 1,481 | 4,788 |
| Recon | 6,325 | 141 | 2,910 | 9,376 |
| KL / OA | 5,919 | 128 | 2,371 | 8,418 |
| MOAKS (O) | 403 | 0 | 194 | 597 |
| MOAKS (C) | 1,414 | 0 | 688 | 2,102 |
| Future OA | 3,385 | 76 | 1,534 | 4,995 |
| Future KR | 6,325 | 141 | 2,910 | 9,376 |

### A. ShapeMed-Knee Dataset Creation

#### 1). Segmentations & Surfaces

We extracted 9,376 Double Echo in Steady State (DESS) knee MRIs from the baseline visit of participants in the OAI [44]. We segmented DESS MRIs automatically using a multi-stage CNN framework; this approach was validated on the OAI dataset, achieving Dice similarity coefficients of 0.99 and 0.91 for femoral bone and cartilage and low ASSD (0.08-0.15mm) [45]. This performance is equivalent to the best-reported cartilage segmentations [6], [33], is the same as expert-human level in terms of cartilage sensitivity to change [46], and is sensitive enough to detect acute changes in cartilage from a 25-minute walking activity [39]. All left knee MRI segmentations were flipped to create right knees and remove variance due to anatomical side. Three-dimensional surfaces were then generated from each femur bone and cartilage segmentation mask using previously established methods [39]; code to create surface meshes is shared for reproducibility.

##### Cartilage Thickness Biomarker

Mean cartilage thickness in pre-defined anatomic regions is a common biomarker for clinical trials and experimental studies [47], [48]. It is critical that NSM-reconstructed surfaces preserve these biomarkers relative to reference surfaces [49]. We calculated cartilage biomarkers with the following processing steps: i) divide cartilage segmentations into subregions, ii) compute cartilage thickness for each vertex over the bone surface, iii) assign each bone-vertex to one of the subregions. Cartilage biomarker calculations used open-source code [50] used in previous investigations [39], [51]. From these data, we computed five cartilage thickness biomarkers as the mean thickness for all bone mesh vertices in each of five established cartilage subregions (trochlea, medial central, lateral central, medial posterior, lateral posterior) [52]. Visualization of cartilage thickness, subregions, and a general orientation to the data are presented in Fig. 2.

**Fig. 2.**
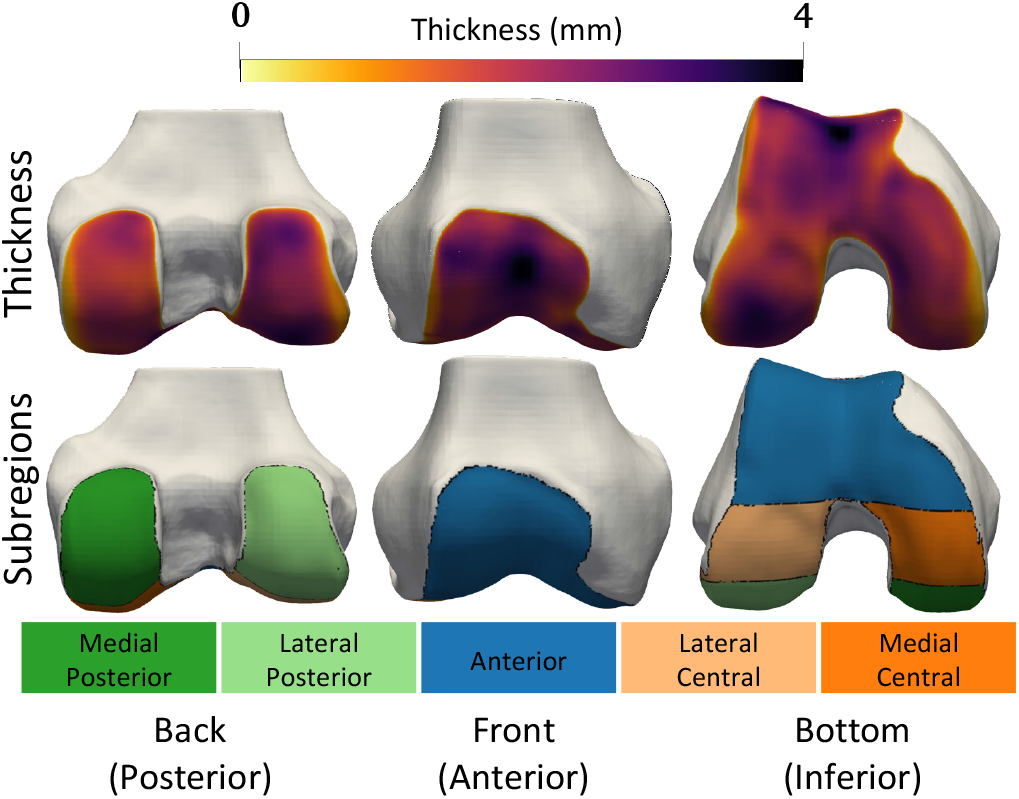
Cartilage thickness (top row) and subregions (bottom row) are displayed on the bone surfaces. Blue is anterior (front), orange is central (middle in front/back axis), green is posterior (back). Dark colors denote medial i.e. the inside of the knee, while light colors denote lateral i.e. the outside.

##### Bone Surface Registration

All femur bones were coregistered to have matching points to create a traditional SSM (Sec. IV) as a baseline model; original full resolution meshes (*∼*220,000 points) were used for the NSMs. First, to reduce the computational complexity of the registration, each bone mesh was downsampled to 20,000 vertices [53], [54]. Next, an average femur shape, determined from 281 knees in a prior study [55], was used as the template and nonrigidly registered to every other bone in the dataset using spectral correspondence-based registration [56], [40] that has been used in multiple knee OA studies [39], [16]. Cartilage thickness and subregions were re-calculated for the registered meshes as described in the previous section Sec. III-C.2. The resulting registered meshes included matching points and cartilage thicknesses for 9,376 femur bones.

##### Mesh Quality Control

To ensure high-quality meshes in the dataset, we generated static images of every bone mesh from 4 orthogonal planes (top, bottom, front, back) using pyVista [57] and an imaging researcher with 10 years of experience with bone analysis manually reviewed every image. From this analysis, we identified 57 meshes (0.6%) with large errors primarily due to physiologically-plausible holes at the sites of anterior cruciate ligament reconstruction. These 57 meshes were removed from the dataset. An additional 9 meshes had moderate errors, and 30 meshes had small potential errors; these meshes were retained in the dataset. IDs for moderate and small error meshes, and quality control images for all knees are provided for dataset users to use their custom exclusion criteria.

### B. Prediction Tasks

OA is a whole joint disease that affects multiple tissues, with an emphasis on the cartilage and bone. We developed five prediction tasks which test a model’s ability to understand shape complexity relevant to current bone and cartilage health as well as future disease progression.

OA is commonly diagnosed using X-rays graded using the Kellgren-Lawrence (KL) system [10]. The KL system assigns knees a grade between 0-4 (0 = no OA, 1 = doubtful OA, 2 = mild OA, 3 = moderate OA, 4 = severe OA). Diagnosis with OA is defined as *KL≥* 2. Beyond diagnosis, KL grading is used in research and clinical trials to “stage” the severity of OA in the whole joint (all tissues/bones) beyond binary classification. Therefore, our first two tasks are:

1. **General OA staging** by predicting KL grade (0-4)
2. **Binary OA diagnosis** (*KL ≥* 2) While KL grading provides a whole-joint OA measure, it is a coarse measurement based on 2D X-rays and does not provide fine-grained, location-specific information in 3D. Therefore, it cannot be used to identify where and what tissues are involved in a person’s disease. The MRI Osteoarthritis Knee Score (MOAKS) measures multiple features of OA that are localized to different regions of the joint [58]. Our third task involves predicting three MOAKS scores (one bone and two cartilage features) in six distinct regions of the femur. MOAKS scoring provides clinically important information and can simultaneously serve as a test of how well a model can spatially localize fine-grained OA features. Task three is:
3. **Advanced localized OA staging** by predicting three MOAKS scores (Score 1: Osteophytes, Score 2: Cartilage Thinning, Score 3: Cartilage Hole) in 6 femoral regions divided across the anterior, central, and posterior regions in the medial and lateral condyles. The three MOAKS scores were defined as follows:
  - ***Score 1 Osteophytes***: Osteophytes are abnormal bone growths (bone spurs) that occur at the edges of the cartilage and are a hallmark sign of OA. The MOAKS osteophyte score includes 4 levels (0: None, 1: small, 2: medium, 3: large). Due to a low prevalence of grade 3 scores (*<* 5%), we binned MOAKS osteophyte score into 3 levels (0-2) where level 2 includes original scores of 2/3.
  - ***Score 2 Cartilage Thinning***: A key sign of OA is cartilage thinning. The MOAKS cartilage thinning score categorizes the % of a region with any cartilage thinning into 4 categories. Given a class imbalance amongst the four categories, we binarize this score as individuals with *<* 10% thinning (grades 0/1) and *>* 10% thinning (grades 2/3). This approach is used in prior OA studies [59].
  - ***Score 3 Cartilage Hole***: The final score quantifies the % of a region that has a full thickness defect (a hole) in the cartilage into the same 4 levels (0-4) as cartilage thinning. Cartilage holes rarely occur (6-16%), thus we binarized this score into no hole (grade 0) and any hole (*≥* 1). The final two tasks were created to test whether a model can predict future OA diagnosis (within 4 years) in currently healthy subjects, and whether a medical event (knee replacement) has occurred (within 9 years). Future OA diagnosis and knee replacement prediction are common tasks performed in the OA literature, are challenging, and would provide valuable information to identify which patients should be treated earlier. MRI-based SSMs of bone shape, and CNN’s applied to X-ray data have previously been used to predict these outcomes [17], [34], [60], [61].
4. **Predict future disease (OA)** within 4 years.
5. **Predict future knee replacement** within 9 years.

### C. Evaluations

#### 1) Surface Reconstruction

We evaluate surface reconstruction errors separately for the bone and cartilage surfaces using ASSD. We test ASSD on the whole test set and separately for the 5 KL grades to assess whether reconstruction errors depend on disease state.

#### 2) Cartilage Thickness Biomarker

To evaluate whether reconstructed bone and cartilage surfaces preserve important cartilage biomarkers, we analyze the five cartilage subregions on the whole test set and on each of the 5 KL grades in the test set. Between the mean thickness of the original and reconstructed surfaces we compute 1) the root mean squared error (RMSE *↓*) to determine absolute errors and 2) the standard deviation of the difference (*SDD ↓*) as a measure of consistency that removes the effect of systematic bias.

#### 3) Prediction Tasks

- ***OA Staging***. OA staging is quantified using the KL grade, a semi-quantitative multi-class measure of OA with variation between raters. As such, relative agreement is commonly used to assess KL predictions and inter-rater agreement. We use accuracy and quadratically-weighted Cohens Kappa, as done previously [62], [63], [64].
- ***OA Diagnosis***. As OA diagnosis is a binary prediction task with relatively well-balanced groups, we compute the common metrics of area under the receiver operating characteristic curve (AUROC) and accuracy.
- ***Advanced OA Staging (MOAKS)***. We assess three MOAKS scores (measuring osteophytes, cartilage thinning, cartilage holes) separately for six regions of interest. Score 1 (osteophytes) includes three classes, and thus we compute quadratically weighted Kappa and accuracy. Since both Score 2 (cartilage thinning) and Score 3 (cartilage hole) are binary tasks with large class imbalance, we compute F1 score and the area under the precision-recall curve (AUPRC).
- ***Future disease (OA)***. The incidence of OA in the four years following baseline was relatively rare, occurring in only 9% of subjects. Therefore, we compute the F1 score and AUPRC.
- ***Future knee replacement surgery***. The incidence of knee replacement in the 9 year follow-up was rare (5%). Therefore, we compute the F1 score and AUPRC.

## IV. Benchmark Models

We compared multiple types of shape models and CNNs on our tasks. We compare an SSM, two implicit NSMs, and our hybrid explicit-implicit NSM for reconstruction tasks. In addition to these models, for the prediction tasks, we also compare 3D CNNs applied to raw image data and to bone/cartilage segmentations. The models are described in the following.

### A Neural Shape Models

DeepSDF-based NSMs train a decoder to take as input a latent vector *z* and coordinate *x* and predict the signed distance *s* of *x*. NSMs typically use an autodecoder framework where *z* is learned by jointly optimizing a dictionary of latents along with the network weights to predict *s* while using regularization so *z* matches a multivariate Gaussian distribution. All NSMs used in this study were trained using the same framework, including point sampling, training hyperparameters, and reconstruction strategy.

#### Point Sampling

Before training, an arbitrary mesh was chosen as the reference. Every other bone mesh was registered to the reference using a similarity transform (rigid + scale); the transform was applied to the coinciding cartilage surface. Next, bone and cartilage meshes were centred using the mean of the bone points and were normalized using maximum radial distance so both tissues lie within a unit sphere. Then, separately for the bone and cartilage surfaces, 500,000 points were sampled. Ninety percent of points were randomly sampled by first sampling positions on the surface using blue noise to produce uniform random samples. Then, sampled surface points were perturbed by adding zero mean Gaussian noise: 45% *σ* = 0.016; 45% *σ* = 0.05. The remaining 10% of points were uniformly sampled over the unit cube. Finally, *s* from both meshes was calculated for every sampled point.

#### Training

Prior to training, each bone/cartilage pair was assigned a random *z∼ 𝒩* (0, 0.01^2^). During training, for each subject (*k*) and surface type (*j*: bone/cartilage), 17,000 points (*X*_*jk*_) were randomly sampled with equal numbers of points inside (-) and outside (+) the surface. Eqn. (1) was optimized to minimize the error in predicted *s* and to regularize the latent *z*. The loss comprises a reconstruction and latent regularization term. The reconstruction term penalizes hard samples (predicted wrong sign) as shown in Eqn (2) and includes a weighted ℒ_1_ where *λ* (0-1) controls the weighting on hard samples with *λ* = 0 being equivalent to regular *ℒ*_1_ and higher values provide greater penalty [24]. *λ* was exponentially increased from 0 to 0.2 over the first 1800 epochs. The latents and network weights *f*_*θ*_ were jointly optimized using the AdamW optimizer with a weight decay of 1e-4 [65]. During training, we used separate learning rates and schedules for the network weights and the latents *z*. For both sets of parameters, learning rate (lr) was decayed as *lr* = *lr*_0_ *×f* ^(*e/i*)^ where *lr*_0_ is the lr at time zero, *f* is the update factor, *e* is the current epoch, and *i* is the interval which *lr* is updated. Network weights had learning parameters of *lr* = 5 *×* 10^*−*3^, *f* = (1*/*1.05), *i* = 16.67 and latents *z* had learning parameters of *lr* = 10^*−*4^, *f* = 0.1, *i* = 1000. A latent regularization loss independently penalized each *z* component with *σ* = 100 to promote a spherical covariance structure. The regularization weight had a linear warmup over the first 100 epochs and was then cyclically annealed with 5 cycles over the training period (2,000 epochs). The cyclic anneal weight *β* for each cycle was defined using Eq. (3) where *t* is the epoch for the current cycle and *T* is the number of epochs in each cycle. We clamped signed distances *s* at |*s*| = 0.1 for the implicit NSM and |*s*| = 1 for the hybrid NSM.

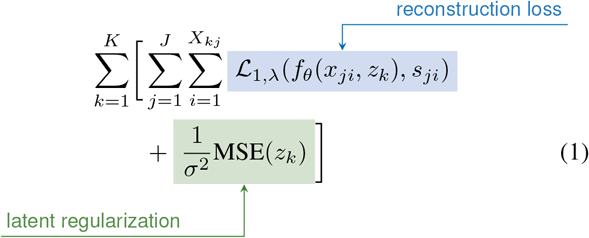

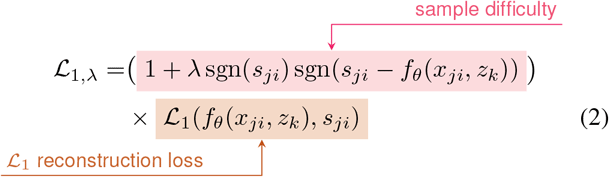

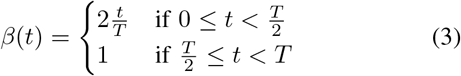

#### Test Time Reconstruction

To reconstruct surfaces and create shape-specific latents, the NSM weights were frozen and the NSM was fit to the new surfaces. Specifically, the bone to be reconstructed was similarity registered to the mean bone shape of the NSM (zero-vector) and the bone/cartilage vertices were scaled to be within a unit sphere. Then, a randomly initialized latent *z∼ 𝒩* (0, 0.01^2^) was optimized for 2,000 epochs to reconstruct the surfaces using an ℒ_1_ loss between the network predicted signed distance *s* and the actual *s* of 20,000 randomly sampled surface points (*s* = 0) using the Adam optimizer. The learning rate was decayed by a factor of 0.9 every 20 epochs, and early stopping was implemented with a patience of 50 epochs. Predicted *s* were clamped at | *s* | = 0.1. No latent regularization was used. To extract the reconstructed surface, a grid of resolution 256 was created over the unit cube; these coordinates and the optimized latent *z* were input into the trained network to predict *s* of each coordinate. The surface was then extracted using marching cubes [66] applied to the coordinates and corresponding *s*. Finally, the reconstructed surface was returned to its initial position and size by applying the inverse of the similarity transform used to initially align the bones.

#### Hybrid Explicit Implicit NSM

The hybrid NSM is based on triplanar architectures [29], [30] as outlined in Fig. 4. A global latent *z* of a length of 512 is processed via a fully connected layer, resulting in a 2048-length vector. This vector is then reshaped to be 2*×* 2 *×*512 before being input into a CNN decoder. The CNN decoder had 5 2D transpose convolution layers, with stride 2 and 512 channels as outputs at each layer. The final output layer of the CNN was sized 64 *×* 64 *×* 384; the 384 features maps were split into 128 features per orthogonal plane. Sampled points *x ∈* ℝ^3^ are projected onto the three orthogonal planes, and a length 128 *z* was obtained per feature plane via bilinear interpolation. Plane features were combined via summation, yielding a length 128 local *z*. The local *z* and the sampled *x* position were concatenated and input into the implicit 3-layer MLP with width 512, *ReLU* activations, and a length two output (one for each tissue) with a tanh activation. Unique from previous work, our model and training pipeline use the triplanar model as an autodecoder only [21], train it using curriculum learning [24] and cyclic annealing of the latent variables [67], and predict multiple output surfaces for a single network.

#### Implicit Multi Layer Perceptron NSM

The implicit decoder was two 8-layer MLPs of width 512, one for each tissue. The same inputs (*x* and *z*) were input into each MLP, and a skip connection was used to input them to layer 4. *ReLU* activations were used throughout. The output of each head was sized one and used the tanh activation.

#### Implicit Modulated Periodic Activation NSM

Implicit decoders with periodic activations have been shown to accelerate learning and improve surface reconstruction [26]. However, generative models did not work directly while using periodic activations. We thus use a Modulated Periodic Activation (MPA) network as proposed previously [25]. Briefly, the MPA uses two networks, a “synthesizer” MLP with sinusoidal activations takes as input the coordinates (*x*) to be queried and a separate “modulator” MLP with *ReLU* activations takes as input the latent variable (*z*). The modulator includes skip connections of the *z* to the input of every layer, and the outputs of each layer of the modulator network are multiplied elementwise by the outputs of the synthesizer network. We used a model depth of 8, and MLP widths of 512 throughout, the same as the traditional implicit MLP. The output was length two (one for each tissue) with a tanh activation.

### B. Statistical Shape Model

The SSM was fit using [50], the same as described in previous investigations [39], [16]. SSM-based reconstruction does not provide explicit cartilage surfaces but instead computes thicknesses at each bone vertex, therefore ASSD was not evaluated for SSM cartilage.

### C. Convolutional Neural Network

We trained two DenseNet121 models as implemented in the MONAI package [68]. One network was trained with an input of the raw DESS MRI data and the other an input of the femur bone and cartilage segmentations. For both variants, the 3D volumes used for input were downsampled from the original volumes (384 *×* 384 *×* 160) to be sized 384 *×* 384 *×* 80, using bilinear interpolation. This approach preserved full-resolution data in-plane, while reducing slice thickness to 1.4mm, which is sufficient for clinical trials including quantitative cartilage analyses [48]. CNNs were trained with a batch size of 8, the AdamW optimizer, an initial learning rate of 10^*−*5^ exponential decay with gamme=0.8 and weight decay=0. Training was performed with a single Nvidia A6000 GPU.

## V. Experiments

### A. Reconstructions

Reconstruction evaluations are provided for NSMs. No reconstruction results are provided for the CNN because it is not generative.

#### Dataset Size

To determine data efficiency, we trained the implicit MLP NSM, hybrid NSM, and SSM models using 4 training set sizes: 50, 200, 1,000, 6,325. NSMs were trained for 2,000 epochs (Tab. V). SSMs were tested using progressively more principal components (Tab. V). These analyses identified that: a) The hybrid NSM performed best for ASSD and both cartilage biomarker measures across dataset sizes, b) Increasing dataset size up to 6,325 increased reconstruction performance for all models, and c) Increasing the number of PCs used in SSM reconstruction did not overfit up to 1,298 PCs (99% explained variance)

The hybrid and the implicit MPA NSMs best reconstructed areas of OA disease (Fig. 3). Fig. 6 distributions of ASSDs in the test set demonstrate that the hybrid NSM had better ASSD for bone (4-17%) and cartilage (8-9%). Tab. II shows that the implicit MPA and the hybrid NSM had the lowest errors for reconstruction and cartilage biomarkers; however, the hybrid NSM performed considerably better in severe disease (KL 4). Better SDD compared to RMSE indicates that all models had a small bias compared to the reference standard (Tab. II).

**TABLE II.**
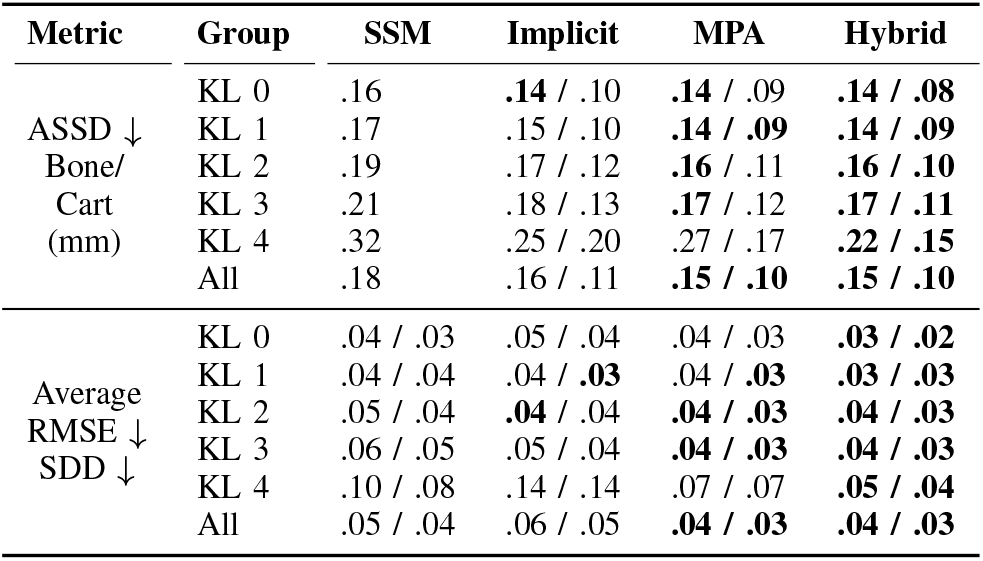
Summary of reconstruction performance for each model (SSM, implicit NSM, hybrid NSM) across the whole test dataset (All) and each KL grade (0-4). Metrics include surface reconstruction errors (ASSD) and cartilage biomarker outcomes (RMSE, SDD) averaged over five regions.

**Fig. 3.**
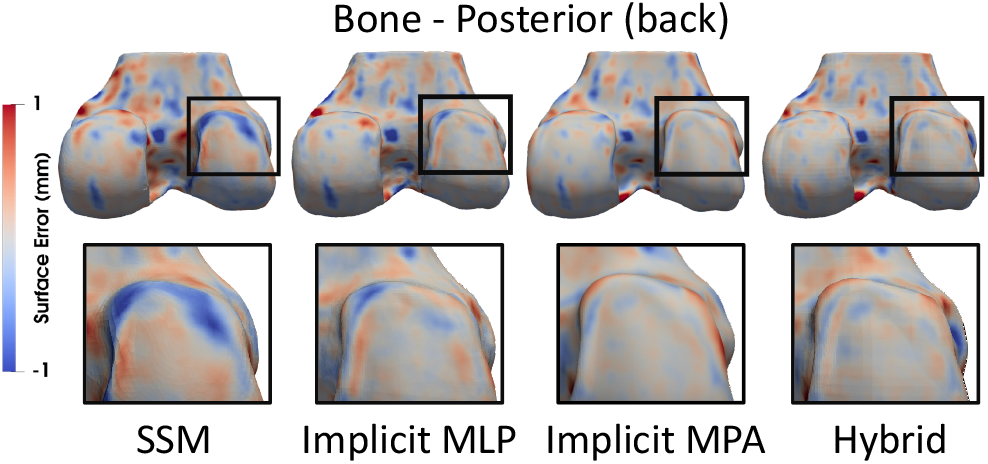
Reconstructed bone and cartilage surfaces colored by reconstruction error. Blue indicates the reconstruction was inside of the reference, and red indicates the reconstruction was outside. Zoomed regions highlight an area of disease (osteophyte on the posterior lateral femur) that was not captured by the SSM (blue), had smaller error for the implicit MLP NSM, and had the least error for the implicit MPA and hybrid NSMs.

**Fig. 4.**
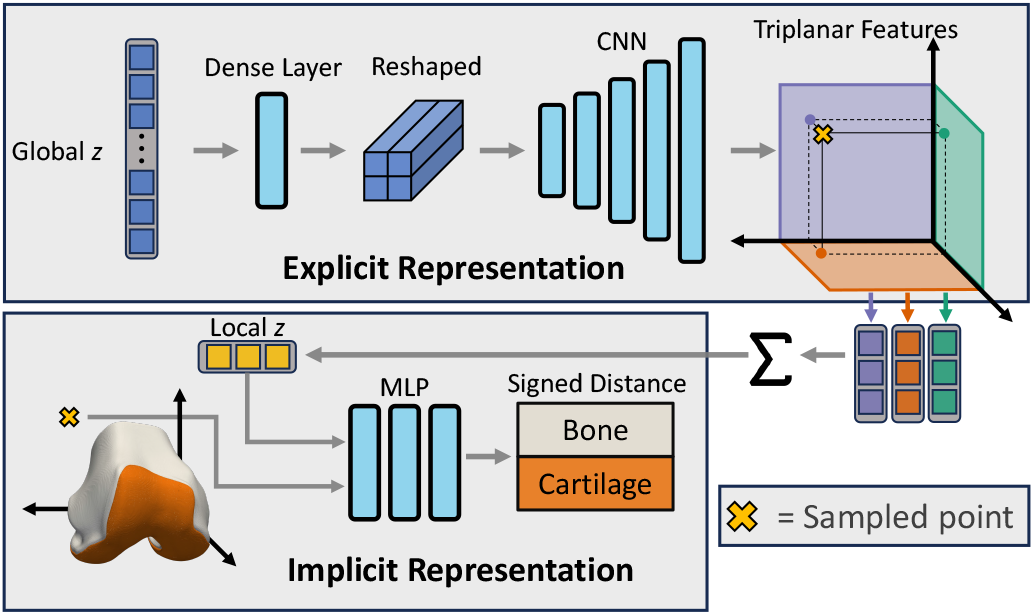
Overview of network architecture. A global latent ***z*** controls the overall generated shape. The global ***z*** is passed through a dense layer, reshaped and then fed through a 5-layer CNN to produce **64 *×* 64** 2D output with 384 feature maps. The 384 feature maps are split into 3 to produce one set of **64 *×* 64 *×* 128** feature maps per orthogonal plane. To determine the signed distance of a particular point (⊗) that point is projected onto each feature map plane, and the corresponding feature vector is extracted using bilinear interpolation. These planespecific feature maps are summed, yielding the local ***z***. The local ***z*** is a coordinate-specific latent vector that controls the signed distance prediction. The local ***z*** along with the XYZ coordinates of point ⊗ are passed to a three-layer multilayer perceptron which outputs the signed distance of the two surfaces (bone and cartilage).

**Fig. 5.**
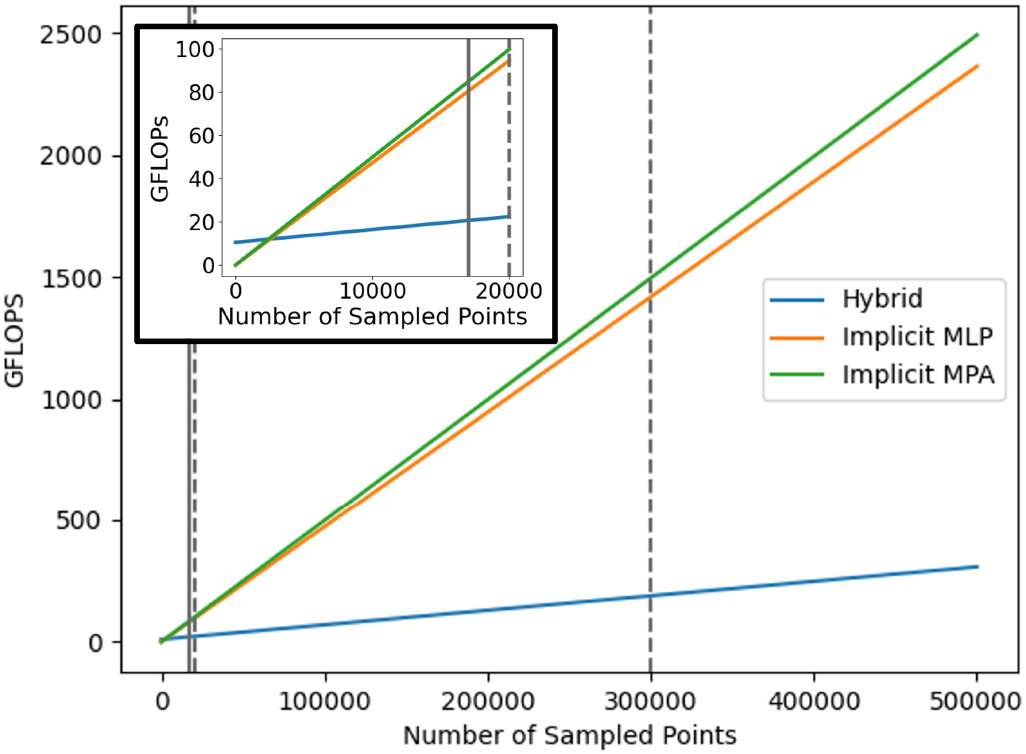
The floating point operations (FLOPs) used by each of the three NSM models for multiple amounts of sampled points. The inset graph shows the plot for relatively small amounts of sampled points of 0-20,000. The vertical lines represent numbers of sampled points used in this study: 17,000 for training, 20,000 for latent optimization during reconstruction, and 300,000 used to reconstruct the original surfaces. The hybrid model has a higher intercept, but a much smaller slope, thus making it more efficient for large, practical, points samples.

**Fig. 6.**
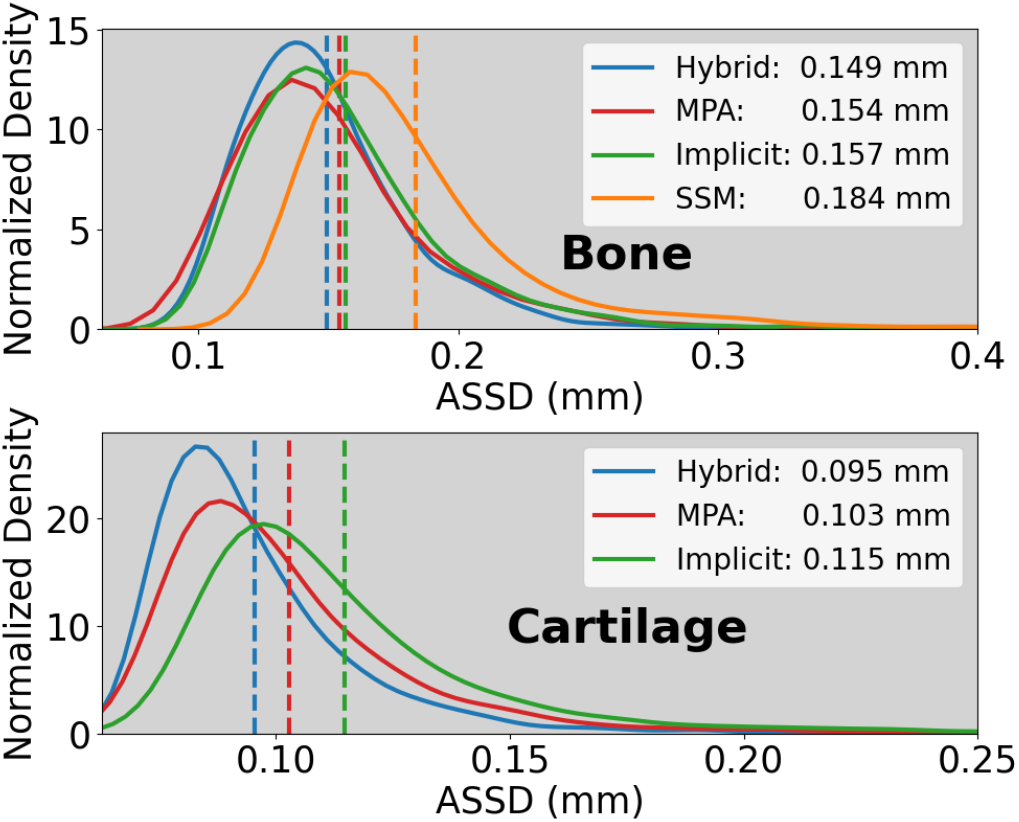
Probability density functions of the bone and cartilage average symmetric surface distances (ASSD). Distribution tails were truncated for visualization purposes.

#### Latent Size

We tested the effect of doubling latent size on ASSD errors for the hybrid and implicit MLP NSMs. Reconstruction accuracy improved as latent size increased, with the hybrid NSM ASSD dropping 26% and 22% for bone and cartilage, respectively (Tab. V).

#### Compute

We compared model compute resources using three metrics. 1. The total parameter count of each model. The number of floating point operations (FLOPs) that each model used for an increasing number of sampled points. The average time it took each model to reconstruct 100 randomly sampled meshes from the test set. The hybrid NSM had the greatest number of parameters (Tab. III), with the two implicit models having roughly equal parameter counts. However, the hybrid model is much more compute efficient, as demonstrated by requiring fewer FLOPs for applicable numbers of sampled points (Fig. 5 and Tab. III) and requiring less time to reconstruct a mesh; the hybrid model was 37% faster vs. the MPA NSM, 16% faster than the MLP NSM. The SSM was fastest.

**TABLE III.** Model information including parameter (PARAM) count, giga (G) floating point operations (FLOPS) for multiple numbers of sampled points (***n***), and the average time needed to reconstruct 100 randomly sampled test set meshes using each network.

| Measures | Hybrid | Implicit MPA | Implicit MLP | SSM |
| --- | --- | --- | --- | --- |
| Param (M) | 20.8 | 5.0 | 4.7 | 3.2 |
| FLOPs (G) $n = 1$ | 10.5 | .005 | .005 | - |
| FLOPs (G) $n = 17,000$ | 20.6 | 84.7 | 80.3 | - |
| FLOPs (G) $n = 20,000$ | 22.4 | 99.7 | 94.5 | - |
| FLOPs (G) $n = 300,000$ | 188.7 | 1,495.0 | 1417.7 | - |
| Recon Time (s) | 33.2 | 52.2 | 39.7 | 20.1 |

### B. Classification / Staging

An MLP was trained to predict each clinical evaluation task using each model’s encoded *z* as input. Hyperparameters were determined via a grid search over depth (2,3), width (64-256), dropout (0.2, 0.4), learning rate (10^*−*3^ to 10^*−*5^), and batchsize (64-512). We also trained two 3D CNNs for clinical prediction tasks Sec. IV-C. Loss functions for CNNs and MLPs included binary cross entropy (OA, MOAKS cartilage thinning and hole, future OA and knee replacement) and consistent rank logits ordinal regression (KL, MOAKS osteophytes) [69].

#### OA Staging & Diagnosis

For predicting KL, the resulting *κ* of the trained models was 0.69-0.79, with the hybrid NSM having the best performance and the implicit MLP NSM having the worst Tab. IV. All models performed comparably to inter-radiologist agreement (0.66-0.89)[63], [70], [71], [64]. Prior X-ray based DL methods performed slightly better (0.83-0.88) [62], [64].

**TABLE IV.** Performance on the prediction tasks using metrics described in Sec. III-C.3. Hybrid NSMS consistently exhibit the best performance. ***κ***: quadratically-weighted kappa; Acc: accuracy; AUROC: area under the receiver operating characteristic curve; AUPRC: area under the precision recall curve; OA: osteoarthritis; KR: knee replacement; LR: logistic regression; MLP: multi layer perceptron; MPA: modulated periodic activations. All shape model predictions used an MLP, except for Hybrid+LR which used LR. Acc and F1 scores for binary tasks (OA, MOAKS hole, MOAKS thinning, Future OA, Future TKR) were optimized on the test set.

| Task | Metric | Method |  |  |  |  |  |  |
| --- | --- | --- | --- | --- | --- | --- | --- | --- |
|  |  | CNN Seg | CNN Image | SSM | MLP | MPA | Hybrid | Hybrid+LR |
| KL | $\kappa$ / Acc | .75 / .59 | .78 / .59 | .78 / .59 | .69 / .54 | .71 / .56 | **.79 / .59** | .72 / **.60** |
| OA | AUROC / Acc | .90 / .84 | .90 / .83 | .91 / .83 | .88 / .81 | .90 / .83 | **.92 / .85** | .92 / .81 |
| MOAKS Osteo | $\kappa$ / Acc | .00 / .49 | .04 / .50 | .16 / .50 | .35 / .54 | .52 / .61 | **.53 / .63** | .46 / .60 |
| MOAKS Cart Thin | AUPRC / F1 | .51 / .56 | .50 / .55 | .64 / .61 | .53 / .54 | .63 / .63 | .74 / **.69** | **.75** / .63 |
| MOAKS Cart Hole | AUPRC / F1 | .32 / .42 | .31 / .41 | .45 / .47 | .44 / .47 | .55 / .55 | **.57 / .58** | .56 / .33 |
| Future OA | AUPRC / F1 | .10 / .19 | **.20 / .29** | .10 / .19 | .15 / .24 | .19 / .26 | .12 / .20 | .14 / .18 |
| Future KR | AUPRC / F1 | .07 / .13 | .29 / .40 | .27 / .33 | .24 / .36 | .30 / .39 | **.33 / .43** | .18 / .27 |

**TABLE V.** Validation set (N=141) reconstruction performance for multiple dataset and latent sizes. There are no average symmetric surface distance (ASSD) results for cartilage reconstruction using the statistical shape model (SSM) because the SSM does not create a cartilage surface. SSM results are for the number of principal components needed to explain 95 and 99% of the variance. NSM: neural shape model.

| Model | Latent size | Dataset size | ASSD Bone | ASSD Cartilage |
| --- | --- | --- | --- | --- |
| Hybrid NSM | 512 | 50 | 0.27 | 0.19 |
|  | 512 | 200 | 0.20 | 0.14 |
|  | 512 | 1,000 | 0.17 | 0.11 |
|  | 512 | 6,325 | 0.15 | 0.09 |
|  | 1,024 | 6,325 | **0.11** | **0.07** |
| Implicit NSM | 512 | 50 | 0.37 | 0.40 |
|  | 512 | 200 | 0.23 | 0.18 |
|  | 512 | 1,000 | 0.17 | 0.13 |
|  | 512 | 6,325 | 0.16 | 0.11 |
|  | 1,024 | 6,325 | 0.13 | 0.09 |
| SSM | 32 (95%) | 50 | 0.58 | - |
|  | 94 (95%) | 200 | 0.42 | - |
|  | 180 (95%) | 1,000 | 0.30 | - |
|  | 269 (95%) | 6,325 | 0.24 | - |
|  | 1298 (99%) | 6,325 | 0.13 | - |

When directly diagnosing OA, the hybrid NSM performed best (AUROC: 0.92) and the implicit MLP NSM performed worst (Tab. IV), similar to the KL task. Interestingly, the CNN applied to the segmentation and the image performed the same, indicating the raw MRI provides no additional information. Accuracy was slightly lower (0.81-0.85) than DL-based Xray OA grading (0.87-0.90) [72], [64]. Our CNN predictions were comparable to a previous CNN applied to MRI data for predicting OA [73].

#### Advanced OA staging

The hybrid NSM performed best for all three MOAKS tasks when averaged over the regions (Tab. IV). These results indicate that the latent *z* fit by the NSM more meaningfully represented both the location and the size of OA features. Not only is this important for OA, but it demonstrates novel capacities of NSMs that are not commonly tested; the ShapeMed-Knee dataset provides a unique method of testing these capacities using real-world data.

The CNN models generally performed worst in predicting MOAKS scores (cartilage holes, cartilage thinning, osteophytes) Tab. IV. Prior DL work uses MOAKS to determine severity of cartilage damage [59]. Other work predicts other features of MOAKS, bone bruises [74] or inflammation [75]. This is the first quantification of MOAKS osteophyte and cartilage health, demonstrating that NSMs encode this important information that is currently prohibitive to obtain clinically, and costly for research and clinical trials.

#### Future OA & knee replacement prediction

All models performed poorly on future event prediction tasks (Tab. IV). The best-performing future OA diagnosis was by the raw image-based CNN (AUPRC: 0.20, F1: 0.29); it is possible non-shape-related features such as bone bruises or joint inflammation boosted CNN image performance [76]. Our general poor future prediction is in contrast to other work using SSMs [17], [35] or CNNs [61], [77] to predict future OA and knee replacement. However, this discrepancy is likely owing to the statistical metrics evaluated, the AUROCs achieved by deep learning papers for future knee replacement were between 0.81 and 0.88 [61], [77], whereas AUROC for our hybrid NSM was 0.87. The overly optimistic results in the literature are likely attributed to the imbalanced data; we suggest future researchers use F1 and AUPRC to provide more balanced results.

### C. Interpretability

One of the powers of shape models is that they are fit in a self-supervised fashion, and are generative. To show the utility of this, we trained a logistic regression classifier on hybrid NSM *z* for each prediction task. Results in Tab. IV show that the simple classifier is one of the best for disease staging. We tested latent interpolation smoothness by assessing the effect of interpolation on reconstructions and disease prediction. Using the hybrid NSM we interpolated *z* from the mean healthy (KL 0) to the mean severe OA (KL 4) shapes in the test set, generated synthetic surfaces, and applied the logistic classifiers on each *z* to determine KL and MOAKS cartilage thinning grades Fig. 7. Shape space interpolation generated smooth physical interpolations and predicted smooth transitions of disease states Fig. 7. This general-purpose representation is powerful because application to other image modalities only requires a segmentation mask, whereas CNN-based approaches would require re-training on entirely new datasets. Furthermore, interpolation could be used to track individual patient disease trajectories over time, opening the door to novel ways of understanding disease.

The generative nature of the NSM enables further validation that classifiers applied to the latent *z* are capturing features of interest. Fig. 8 takes the latent *z* fitted to a patient, and interpolates it along the vector defined by a logistic regression classifier that predicts medial cartilage holes. Simple linear interpolation along the fitted vector precisely controls the size of the cartilage hole on the medial side. This visualization improves confidence in the fitted model, but may also enable entirely new applications. For example, it is possible to precisely add and remove specific, localized, features of disease and therefore to generate synthetic versions of a patient’s anatomy. These synthetic digital twins can be used for *in silico* simulations to determine the effects of specific disease features on tissue biomechanics [14], or to inform surgical planning such as cartilage repair [78], [79]. Importantly, this example uses simple linear interpolation; future work can leverage latent diffusion models [80] to advance this capacity.

## VI. Discussion

The developed ShapeMed-Knee dataset enabled us to train, test, and compare 5 models, including 4 types of shape model, and a CNN. The included reconstruction and clinical prediction tasks highlighted that although a model may be better for reconstruction (implicit MPA vs implicit MLP; Tab. II), it may perform worse for prediction tasks, such as cartilage localization (Tab. IV). This result highlights the importance of creating medical-specific datasets and evaluations. We envision the community leveraging this dataset, its pre-defined data splits, and evaluation criteria to develop and evaluate new and better models specific to the medical domain.

The proposed hybrid NSM performed best for all reconstruction tasks, both conventional and cartilage biomarkerrelated (Tab. II). The hybrid NSM performed best for all clinical prediction tasks except future OA prediction. As a whole, the hybrid NSM provided a single model that is capable of high reconstruction accuracy and a broad array of clinically relevant prediction tasks. While the hybrid NSM performed best at reconstructing diseased joints (KL 4), it still performed worse than for healthier joints (KL 0-3). Future work should explore methods to promote the reconstruction of disease features, such as using multi-task learning to explicitly predict OA features such as osteophytes or cartilage holes while training the NSM. Future work should also work on modeling a greater number of inter-connected tissues, making hybrid NSMs even more efficient, and integrating latent diffusion for latent interpolation.

A major strength of shape models is that they are trained in a self-supervised manner. That is, each shape model is first trained on self-reconstruction, creating a latent space that semantically encodes shape information. Performing downstream tasks can then be achieved by training relatively simple models, in our case either an MLP or logistic regression classifier (Tab. IV), to predict the output of interest using latent codes fit to new data using the shape model. In the current investigation, the same latent representations were used for all 5 clinical prediction tasks and outperformed taskspecific CNNs for the majority of clinical tasks. Additional benefits of using a generative self-supervised approach is that interpolating latent space can be used as explainable features of the learned shape space and classifiers trained on the latents (Figs. 7 and 8). Importantly, the shape model can be trained on large datasets, such as the OAI used in this study, and then researchers can use latents fit by this model to train new predictors on smaller available datasets, as was done for many of the clinical prediction tasks. Finally, these shape models can be used on other modalities, such as computed tomography, that can produce a segmentation.

**Fig. 7.**
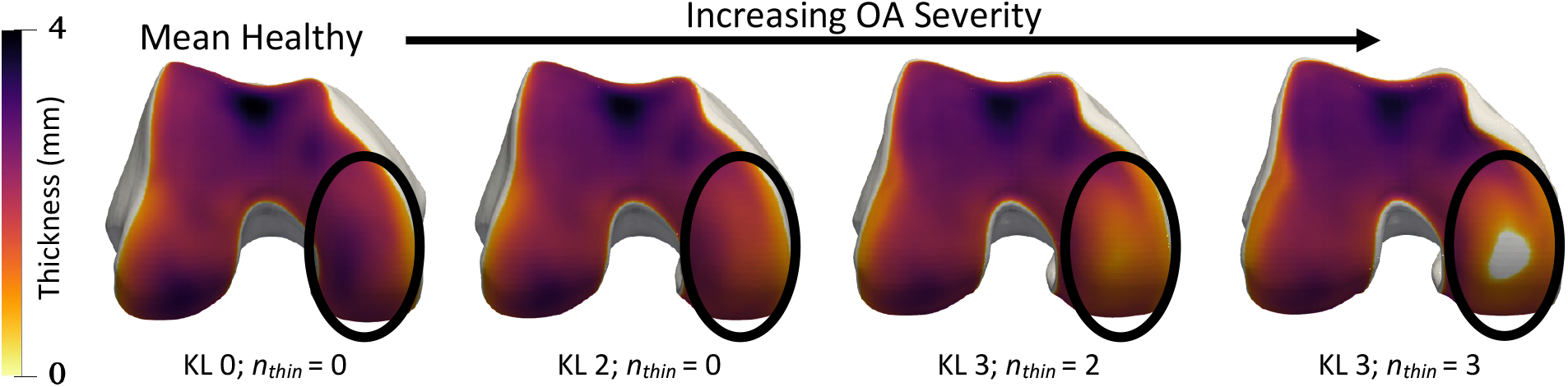
Interpolation in hybrid NSM shape space along the mean healthy to the mean severe OA axis. Smooth progression of cartilage thinning occurs on the medial central femur (circled) with a hole (grey) occurring at the end. Each bone is annotated with disease stage classifications determined by logistic regressions, KL grade, and the number of regions with cartilage thinning (***n***_thin_).

**Fig. 8.**
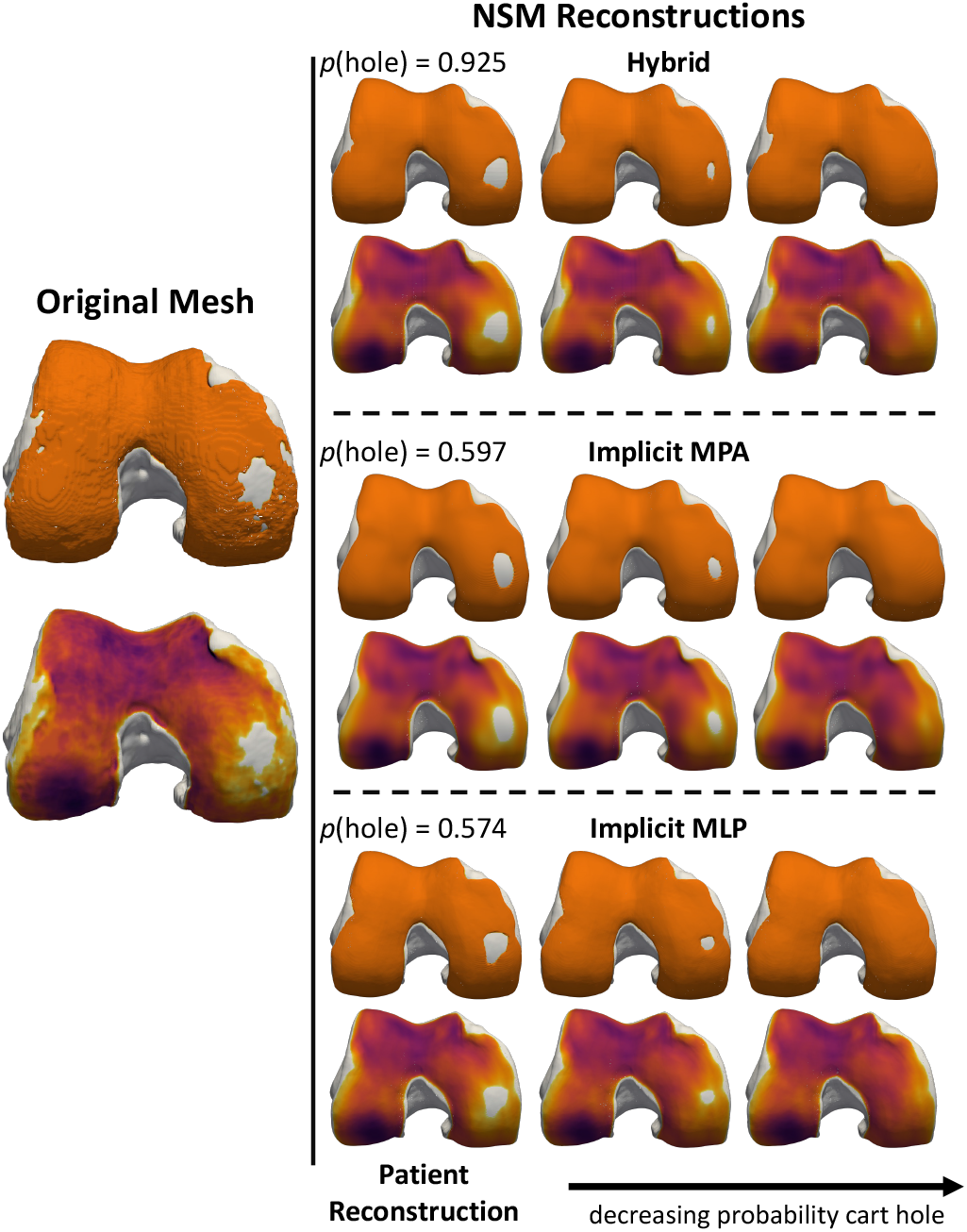
Interpretation of logistic regression-based MRI Osteoarthritis Knee Score (MOAKS) medial cartilage hole classifiers. Each bone is showed as a pair, with the top bone showing the solid cartilage surface in orange and the bottom bone showing the thickness map with the cartilage removed. The left pair is of the original mesh. The right column includes reconstructions derived from each of the NSMs. Within the reconstructions, the left set of meshes is the NSM reconstruction of the patient and shows that each model captures a slight variation of the cartilage hole; using the classifier, the probability of there being a hole (***p*(*hole*)**) obtained from the logistic regression classifier is printed on top of each reconstruction. The other two columns are synthetic bone and cartilage surfaces generated by interpolating the patient-fitted latent ***z*** along a vector defined by the logistic regression coefficients. The synthetic bones progressively close the cartilage hole, while generally leaving the other bone and cartilage surfaces the same. Specific control of anatomical features indicates that these features can be monitored longitudinally and that synthetic alternatives to patient anatomy can be generated for *in silico* simulations.

For OA diagnosis, all models performed comparably to inter-radiologist agreement (0.66-0.89)[63], [70], [71], [64]. However, accuracy was slightly lower (0.81-0.83) than DL models applied to X-rays for OA diagnosis (0.87-0.90) [72], [64]. This finding is not unexpected; first, only femur bones were used by the shape models and KL grade used for diagnosis is performed on the whole joint. Furthermore, KL grading and OA diagnosis are performed using 2D X-rays, and we expect that using the same data used for original grading would yield better agreement. This is particularly true because it is well established that parallax effects in 2D X-rays lead to errors; for example, KL grading can change due to small (5^*o*^) changes in knee flexion angle [9]. Thus, the X-ray based KL grade and OA diagnosis may not be the “true” grade. While the ground truth for this study is the KL grade and OA diagnosis from X-rays, the 3D models do not suffer from parallax effects and thus by training over thousands of examples may produce more accurate predictions. Future work should explicitly test this hypothesis and study how 3D data may be used to overcome parallax effects of X-ray based KL grades.

The emphasis of this study is NSMs. However, we in-cluded CNNs for the clinical prediction tasks to evaluate how NSMs fair compared to these more commonly used models. CNNs performed comparably to the shape models for OA staging and classification. These tasks were the easiest ones proposed in the clinical evaluation, were the most balanced, and had the greatest amount of training data. The CNNs performed particularly poorly for the MOAKS tasks, this is likely partly explained by the smaller amount of available data to train the classifiers for these tasks, and the subtlety of the MOAKS features. This is because the shape models are pre-trained on all training data, learning the distribution of 3D anatomic shapes from a large dataset and organizing it in a continuous latent space. Classifiers are then trained on the organized latents fit using the shape models. This ability to train prediction models using limited data is a benefit of shape models broadly. However, CNNs applied to the raw MR images directly performed best for future OA prediction. This is likely due to a few factors: i) the shape model only includes explicitly encoded information, e.g., femur bone and cartilage, and thus misses other structures like the tibia bone, or ligaments and tendons, ii) the raw image data provides additional information, such as bone texture indicative of bone bruises, or fluid in the knee indicative of overall swelling. This additional information may have enabled better performance for these future prediction tasks.

We presented three classes of NSM: implicit MLP, implicit MPA, and a hybrid explicit implicit NSM. The hybrid model performed the best of these models for nearly every recon-struction and clinical evaluation task. The hybrid model is the largest (20.8 M vs *≤* 5 M params). However, the hybrid model includes two stages and thus it does not use the whole network capacity for every coordinate sampled. As shown in Fig. 4, the hybrid model passes a global *z* through the CNN once, and then uses a much smaller MLP to predict individual coordinate signed distances. As such, when analyzing practical numbers of sampled points either for training, latent optimization, or surface reconstruction, the implicit MLP and implicit MPA models use 3.9 *−* 7.9 *×* more FLOPs Tab. III. To further test this efficiency, we timed the reconstruction of 100 randomly sampled meshes from the test set, and the hybrid NSM had an average reconstruction time 37% faster than the implicit MPA and 16% faster than the implicit MLP; the conventional SSM is still fastest, with reconstructions taking 20.1 s vs 33.2 s for the hybrid NSM. Therefore, the hybrid NSM provides a means of encoding rich shape information in a relatively dense explicit CNN, but leverages a lightweight MLP for efficiency. It is also interesting to compare the MLP (a simple feed forward network) with the MPA (a base MLP with periodic activations whose layer outputs are modulated by a second ReLU-based MLP). This MPA formulation outperforms an MLP that is of the same parameter count, pointing to efficiencies provided by periodic activations.

## VII. Conclusion

We contribute a hybrid explicit-implicit NSM pipeline which demonstrates state-of-the-art performance for multitissue anatomic reconstruction, and clinical outcome prediction. Model training and evaluation were enabled by our new ShapeMed-Knee dataset. All shape models were capable of simple OA staging. Hybrid and MPA NSMs quantified the location and size of OA features for the first time. While hybrid NSMs provide current state-of-the-art bone and cartilage reconstruction, further advances applied to our ShapeMed-Knee dataset have the potential to improve results and, in turn, our understanding of OA. Future work should expand upon our ShapeMed-Knee dataset to include a more complete set of musculoskeletal anatomies, starting with the tibia and patella bones of the knee. Research should also be focused on accelerating the fidelity and speed at which we can reconstruct multiple inter-related anatomic surfaces. We encourage the community to leverage ShapeMed-Knee data and benchmarks to tackle the unique challenges presented by modeling multiple anatomic surfaces and encoding meaningful disease-specific information.

## Data Availability

All data produced are available online at https://huggingface.co/datasets/aagatti/ShapeMedKnee

## Footnotes

1 https://huggingface.co/datasets/aagatti/ShapeMedKnee

2 https://huggingface.co/aagatti/ShapeMedKnee

3 https://github.com/gattia/nsm

4 https://github.com/gattia/shapemedknee

5 https://huggingface.co/datasets/aagatti/ShapeMedKnee

